# Association between circulating inflammatory markers and adult cancer risk: a Mendelian randomization analysis

**DOI:** 10.1101/2023.05.04.23289196

**Authors:** James Yarmolinsky, Jamie W Robinson, Daniela Mariosa, Ville Karhunen, Jian Huang, Niki Dimou, Neil Murphy, Kimberley Burrows, Emmanouil Bouras, Karl Smith-Byrne, Sarah J Lewis, Tessel E Galesloot, Lambertus A Kiemeney, Sita Vermeulen, Paul Martin, Demetrius Albanes, Lifang Hou, Polly A Newcomb, Emily White, Alicja Wolk, Anna H Wu, Loïc Le Marchand, Amanda I Phipps, Daniel D Buchanan, the International Lung Cancer Consortium, the PRACTICAL consortium, Sizheng Steven Zhao, Dipender Gill, Stephen J Chanock, Mark P Purdue, George Davey Smith, Paul Brennan, Karl-Heinz Herzig, Marjo-Riitta Jarvelin, Abbas Dehghan, Mattias Johansson, Marc J Gunter, Kostas K Tsilidis, Richard M Martin

## Abstract

**Background:** Tumour-promoting inflammation is a “hallmark” of cancer and conventional epidemiological studies have reported links between various inflammatory markers and cancer risk. The causal nature of these relationships and, thus, the suitability of these markers as intervention targets for cancer prevention is unclear.

**Methods:** We meta-analysed 6 genome-wide association studies of circulating inflammatory markers comprising 59,969 participants of European ancestry. We then used combined *cis*-Mendelian randomization and colocalisation analysis to evaluate the causal role of 66 circulating inflammatory markers in risk of 30 adult cancers in 338,162 cancer cases and up to 824,556 controls. Genetic instruments for inflammatory markers were constructed using genome-wide significant (*P* < 5.0 x 10^-8^) *cis*-acting SNPs (i.e. in or ±250 kb from the gene encoding the relevant protein) in weak linkage disequilibrium (LD, r^2^ < 0.10). Effect estimates were generated using inverse-variance weighted random-effects models and standard errors were inflated to account for weak LD between variants with reference to the 1000 Genomes Phase 3 CEU panel. A false discovery rate (FDR)-corrected *P*-value (“*q*-value”) < 0.05 was used as a threshold to define “strong evidence” to support associations and 0.05 ≤ *q*-value < 0.20 to define “suggestive evidence”. A colocalisation posterior probability (PPH_4_) > 70% was employed to indicate support for shared causal variants across inflammatory markers and cancer outcomes.

**Results:** We found strong evidence to support an association of genetically-proxied circulating pro-adrenomedullin concentrations with increased breast cancer risk (OR 1.19, 95% CI 1.10-1.29, *q*-value=0.033, PPH_4_=84.3%) and suggestive evidence to support associations of interleukin-23 receptor concentrations with increased pancreatic cancer risk (OR 1.42, 95% CI 1.20-1.69, *q*-value=0.055, PPH_4_=73.9%), prothrombin concentrations with decreased basal cell carcinoma risk (OR 0.66, 95% CI 0.53-0.81, *q*-value=0.067, PPH_4_=81.8%), macrophage migration inhibitory factor concentrations with increased bladder cancer risk (OR 1.14, 95% CI 1.05-1.23, *q*-value=0.072, PPH_4_=76.1%), and interleukin-1 receptor-like 1 concentrations with decreased triple-negative breast cancer risk (OR 0.92, 95% CI 0.88-0.97, *q*-value=0.15), PPH_4_=85.6%). For 22 of 30 cancer outcomes examined, there was little evidence (*q*-value ≥ 0.20) that any of the 66 circulating inflammatory markers examined were associated with cancer risk.

**Conclusion:** Our comprehensive joint Mendelian randomization and colocalisation analysis of the role of circulating inflammatory markers in cancer risk identified potential roles for 5 circulating inflammatory markers in risk of 5 site-specific cancers. Contrary to reports from some prior conventional epidemiological studies, we found little evidence of association of circulating inflammatory markers with the majority of site-specific cancers evaluated.

## Background

Emerging evidence implicates chronic inflammation in cancer development [1–3]. Preclinical studies have shown that pro-inflammatory cytokines (e.g. tumour necrosis factor-α, interleukin-1, interleukin-6) promote cancer cell proliferation, invasion, and metastasis, and transcription factors for these markers (e.g. NF-kB and STAT3) are up-regulated across most cancers [4–8]. Prospective observational studies have reported associations between circulating inflammatory markers and risk of cancer across various anatomical sites [9–23]. Further, pharmacological inhibition of key inflammatory mediators (e.g. COX enzymes, interleukin-1β) in clinical trials has led to reduced risk of site-specific cancers [24, 25]. These successful trial results suggest that pharmacological targeting of other inflammatory markers identified in the observational epidemiological literature could be an effective approach for cancer prevention [26].

However, there are important challenges that accompany the translation of findings from observational studies into effective cancer control strategies. This is because of the susceptibility of conventional observational designs to various biases such as residual confounding (e.g. due to unmeasured or imprecisely measured confounders) and reverse causation [27, 28]. These biases frequently persist despite statistical and methodological efforts to address them [29–31], making it difficult for observational studies to reliably conclude that a risk factor is causal, and thus a potentially effective intervention target [32].

Mendelian randomization (MR) uses germline genetic variants as instruments (“proxies”) for risk factors to generate estimates of the effects of these factors on disease outcomes in observational settings [32, 33]. Since germline genetic variants are quasi-randomly assorted at meiosis and are fixed at conception, MR analyses should be less susceptible to conventional issues of confounding and cannot be influenced by reverse causation bias. In addition, MR analysis considers the long-term effect of risk factors on health outcomes, which is relevant in the context of diseases like cancer where there may be long induction periods between exposure to a particular risk factor and disease initiation [34].

Previous MR analyses that have examined the association of circulating inflammatory markers with cancer risk have been restricted to examining single inflammatory markers [35–41], individual cancer sites [36, 37, 42, 43], or have evaluated the effects of specific classes of inflammatory markers (i.e. cytokines) [44–46]. To date, however, no studies have used a systematic approach to comprehensively evaluate different classes of circulating inflammatory markers across adult cancers.

We aimed to systematically evaluate the causal relationship of circulating inflammatory markers with risk of 30 adult cancers. First, we performed a meta-analysis of genome-wide association studies (GWAS) of circulating inflammatory markers to generate novel and stronger genetic instruments for these markers. Second, we used the Open Targets Platform to identify inflammatory markers with prior evidence from preclinical and/or epidemiological studies to support their aetiological role in site-specific cancers and tested relationships of these inflammatory marker-cancer pairs using combined Mendelian randomization and colocalisation analysis (“Validation analyses”). Third, for all remaining inflammatory-marker cancer pairs, we systematically tested their relationship using combined Mendelian randomization and colocalisation analysis to identify potential novel circulating inflammatory markers implicated in cancer risk (“Discovery analyses”).

## Methods

### Identification of inflammatory markers

We compiled a list of all inflammatory markers that corresponded to one or more of the following classes: acute phase proteins, chemokines, growth factors, interferons, interleukins, and tumour necrosis factors [47–53]. Inflammatory markers were then mapped to their UniProt ID, resulting in 218 unique markers [54].

### Identification of GWAS for inclusion in meta-analysis

GWAS of circulating inflammatory markers were included in meta-analyses if they met the following criteria: i) the study was performed in individuals of European ancestry, ii) the study was adjusted for basic covariates only (e.g. age, sex, principal components of genetic ancestry) to avoid issues due to potential collider bias, iii) effect estimates were presented in standard deviation (SD) units or equivalent (e.g. protein concentrations were inverse-normal rank transformed), and iv) complete summary genetic association data were available for *cis*-acting SNPs (i.e. ±250 kb from the gene encoding each marker). Where two studies with > 50% sample overlap measured the same protein, we selected the larger study for inclusion into the meta-analysis. In total, 6 studies met all inclusion criteria and 8 were excluded [55–60]. A summary of each included study is presented in **Table 1** and a list of excluded studies along with their justification for exclusion is presented in **Supplementary Table 1.**

**Table 1.** Summary of studies included in GWAS meta-analysis of circulating inflammatory markers.

| **Study** | **N proteins** | **Participants** | **Units** | **Adjustment** | **Protein assay** |
| --- | --- | --- | --- | --- | --- |
| Karhunen et al. [58] | 47 inflammatory proteins | 840-13,365 Finnish | Inverse-normal rank transformation | Age, sex, first 10 genetic principal components | Bio-Rad Bio-plex assays, custom panel |
| Folkersen et al. [55] | 90 proteins | 30,931 European | NPX values of proteins (on the log2 scale) were rank-based inverse normal transformed and/or standardised to unit variance | Variable across studies | Olink |
| Gilly et al. [56] | 257 proteins | 1,328 Greek | Inverse-normal transformation of the residuals | Age, age<sup>2</sup>, sex, plate number, per-sample mean NPX value across all assays. Adjustment for season. | Olink |
| Hillary et al. [57] | 70 inflammatory proteins | 1,936 European older adults | Standardised residuals from these regression models were brought forward for all genetic-protein and epigenetic-protein analyses. | Age, sex, four genetic principal components of ancestry, array plate | Olink |
| Pietzner et al. [59] | 179 proteins | 10,708 European | Rank-based inverse normal transformation | Age, sex, sample collection site, 10 principal components | SomaScan |
| Sun et al. [60] | 2,995 proteins | 3,600 European | Rank-inverse normalized | Age, sex, duration between blood draw and processing, first three principal components of ancestry from multi-dimensional scaling | SomaScan |

### Data pre-processing and quality control

For each inflammatory marker of interest, UniProt IDs were mapped to proteomic platform-specific IDs based on annotations provided by platform vendors and manual review [61]. In quality control, 14 markers with the following issues were flagged and removed from one or more studies: those with ambiguous or duplicate Uniprot IDs, unique Uniprot IDs with duplicate probes, genes located on chromosome X, or proteins where summary genetic association data could not be accessed from the relevant data repository (**Supplementary Table 2**). After removal of problematic inflammatory markers, genetic association data for 204 of 218 markers of interest were available in at least one study. For each of these markers, the relevant protein-coding gene for the marker was identified using the UniProt ID mapping function and genomic coordinates for the gene (build GRCh37) were extracted using BioMart [62]. Of the 204 inflammatory markers, 116 markers were only measured in one study with the remaining 88 markers taken forward to meta-analysis.

### GWAS meta-analysis to develop genetic instruments for inflammatory markers

Across all studies, summary genetic association data for each marker were extracted for *cis*-acting variants (i.e. ±250 kb from the gene encoding the protein). Genomic coordinates in Gilly et al. were converted from build GRCh38 to GRCh37 prior to data extraction using LiftOver [63]. All SNPs with a minor allele frequency (MAF) < 0.01 and all palindromic SNPs with a MAF > 0.40 were removed. For inflammatory markers measured in both Sun et al. and Folkersen et al., summary genetic association data from Sun et al. were not included in meta-analyses due to participant overlap across studies [55, 60]. Meta-analyses across inflammatory markers were performed using inverse variance-weighted fixed-effects models in METAL [64]. Of the 88 inflammatory markers included in the meta-analysis, 45 had 1 or more genome-wide significant (*P* < 5.0 x 10^-8^) variants associated with that marker and, therefore, were included in subsequent MR analyses.

### Pair-wise comparison of SNP effects across studies

To compare agreement of SNP effects across studies included in meta-analyses, standardised effect estimates for inflammatory markers were systematically compared across studies using Pearson correlation coefficients. These analyses were performed by extracting independent *cis*-acting SNPs (r^2^<0.01, with reference to the 1000 Genomes Phase 3 CEU Panel) across two *P*-value thresholds (*P* < 5.0 x 10^-8^, *P* < 5.0 x 10^-4^) in PLINK. Pair-wise comparisons were not performed for Folkersen et al. and Sun et al. due to participant overlap across studies. Comparisons were performed by aligning effect directions of SNPs to the protein-increasing allele to prevent inflated Pearson correlation coefficients [65]. The median (interquartile range, IQR) agreement in study-level comparison was r=0.66 (0.24-0.84) when using a *P* < 5.0 x 10^-8^ threshold and r=0.65 (0.45-0.85) when using a *P* < 5.0 x 10^-4^ threshold.

### Genetic instrument construction

Following meta-analysis, genome-wide significant (*P* < 5.0 x 10^-8^) SNPs for each inflammatory marker of interest were extracted and SNPs with evidence of heterogeneity of effect across studies (P*_het_* < 0.001) were removed. In total, 45 inflammatory markers included in meta-analyses were retained and combined with 21 markers measured in a single study (i.e. not included in the meta-analysis) that had 1 or more *cis*-acting genome-wide significant (*P* < 5.0 x 10^-8^) variant associated with the marker [66]. Genetic instruments to proxy 66 circulating inflammatory markers were constructed from SNPs that were permitted to be in weak linkage disequilibrium (LD, r^2^ < 0.10), increasing the proportion of variance in each marker explained by the instrument and, thus, maximising instrument strength [67].

### Cancer GWAS study populations

We obtained summary genetic association data from GWAS of 30 cancer outcomes representing 12 anatomical sites and 18 cancer subtypes within these sites [68–78]. The median (IQR) number of cases across GWAS of unique anatomical sites was 15,161 (8,288-33,604). Analyses in each study were restricted to individuals of European ancestry. Further information on statistical analysis, imputation, and quality control measures for these studies is available in the original publications. A summary of the numbers of cases and controls across each cancer outcome is presented in **Supplementary Table 3**.

### Analytical approach

We employed a two-stage approach to evaluate the effect of circulating inflammatory markers on cancer risk. We first attempted to validate previously reported inflammatory marker-cancer associations from the preclinical and/or epidemiological literature using the Open Targets platform (“Validation analyses”)[79]. The Open Targets platform integrates data (e.g. gene expression, animal models, text mining, pathways and systems biology) from > 20 public sources and uses this data to systematically build an “Overall association score” between drug targets and disease outcomes (i.e. a summary of the overall evidence implicating a protein-coding gene in a disease outcome). All inflammatory marker-cancer pairs with an “Overall association score” ≥ 0.05 were included in validation analyses (scores range from 0-1, where 1 represents strong evidence that a protein is implicated in a disease outcome). Of all remaining inflammatory marker-cancer pairs not included in “Validation analyses”, we then performed a “hypothesis-free” pan-cancer assessment to identify potential novel inflammatory marker-cancer associations (termed “Discovery analyses”).

Mendelian randomization can generate unbiased estimates of causal effects of exposures on disease outcomes if the following assumptions are met: i) the instrument is strongly associated with the exposure (“relevance”), ii) there are no common causes of the instrument and outcome (“exchangeability”), and iii) there is no direct effect of the instrument on the outcome (“exclusion restriction”). Under the assumption of monotonicity (i.e. the direction of the effect of the instrument on the exposure is consistent across all individuals), MR can provide valid point estimates for those participants whose exposure is influenced by the instrument (i.e. complier average causal effect)[80].

For inflammatory markers instrumented by a single SNP, the Wald ratio was used to generate effect estimates and the delta method was used to approximate standard errors. For markers instrumented by two or more SNPs, inverse-variance weighted (IVW) random-effects models were used to estimate causal effects [81]. Standard errors from IVW models were inflated to account for weak linkage disequilibrium between SNPs by incorporating a correlation matrix using the 1000 Genomes Phase 3 CEU reference panel [82, 83].

We evaluated the “relevance” assumption by generating estimates of the proportion of variance in each inflammatory marker explained by the instrument (r^2^) and F-statistics. An F-statistic > 10 is conventionally used to indicate that instruments are unlikely to suffer from weak instrument bias [84]. Colocalisation analysis was performed to evaluate whether circulating inflammatory markers and cancer outcomes shared the same causal variant within a locus, necessary but not sufficient to infer causality between these traits. Such an analysis can also permit evaluation of whether circulating inflammatory markers and cancer endpoints are influenced by distinct causal variants that are in linkage disequilibrium with each other, indicative of horizontal pleiotropy (an instrument influencing an outcome through pathways independent to that of the exposure), a violation of the exclusion restriction assumption. Colocalisation analysis was performed by generating ± 250 kb windows from the sentinel SNP used to proxy each inflammatory marker. We employed a colocalisation posterior probability (PPH_4_) of > 0.70 to indicate support for shared causal variants across circulating inflammatory markers and cancer outcomes. All colocalisation analyses were performed using GCTA-COJO and the coloc package as implemented in Pair-Wise Conditional analysis and Colocalisation analysis (PWCoCo)[85–87]. We used default prior probabilities that any SNP within the colocalisation window was associated exclusively with inflammatory marker concentrations (p_1_ = 1 x 10^-4^), exclusively with cancer risk (p_2_ = 1 x 10^-4^), or both traits (p_12_ = 1 x 10^-5^). Finally, iterative leave-one-out analysis was performed iteratively removing one SNP at a time from multi-SNP instruments to examine whether findings were driven by a single influential SNP.

To account for multiple testing, a Benjamini-Hochberg false discovery rate (FDR) correction was applied across “Validation” and “Discovery” analyses separately [88]. We used an FDR-corrected *P*-value (termed “*q-*value”) threshold of < 0.05 to define “strong evidence” to support analyses, with findings between *q*-value ≥ 0.05 and *q*-value < 0.20 defined as “suggestive evidence”.

### Sensitivity analyses to explore potential aptamer or epitope binding effects

Genetic instruments may be associated with circulating protein concentrations due to aptamer or epitope binding artefacts when using protein assays that rely on binding (e.g. SomaScan)[89]. SNP associations in GWAS that employ aptamer-based protein platforms may therefore represent associations with protein measures due to differential binding rather than differences in protein abundance. As variants sensitive to aptamer or epitope binding effects tend to be missense variants, for two studies that used the aptamer-based SomaScan assay (i.e. Sun et al., Pietzner et al.), we flagged all instruments that were missense variants or variants that were in high LD (r^2^ > 0.80) with a missense variant using functional consequence data from the Open Targets platform and the LDlinkR package [59, 60, 79, 90]. For top findings from “Validation” and “Discovery” analyses generated using multi-SNP instruments, as sensitivity analyses we then re-calculated MR estimates dropping missense variants or variants in high LD with missense variants from instruments. For top findings consisting of single-SNP instruments that were missense variants or in high LD with a missense variant, we explored whether these instruments were also expression quantitative trait loci (eQTL) or splicing quantitative trait loci (sQTL) for the gene encoding the relevant inflammatory marker in the Genotype-Tissue Expression (GTEx) project V8. If missense variants (or variants in high LD with missense variants) also influence expression or alternative splicing of pre-mRNA of the gene encoding the relevant inflammatory marker, causal inference using these variants as instruments is unlikely to be biased even if effect estimates are invalid [91].

### Expression quantitative trait loci enrichment to examine tissue-specific regulatory mechanisms of effects

For all top findings from “Discovery” and “Validation” analyses we examined whether instruments overlapped with eQTL (*P* < 5.0 x 10^-8^) to identify potential tissue-specific regulatory effects of instruments using data on 15,201 RNA-sequencing samples from 49 tissues of 838 post-mortem donors in the GTEx project V8 and 1,544 RNA-sequencing samples from 13 immune cells of 91 healthy subjects in the Database for Immune Cell Expression (DICE) [92, 93]. Where there was eQTL overlap, we then used multiple trait colocalisation (“moloc”) to evaluate colocalisation across circulating inflammatory marker concentrations, tissue-specific or immune cell-specific gene expression, and cancer risk [94]. We employed a colocalisation posterior probability > 0.70 to indicate support for shared causal variants across all three traits. We used default prior probabilities that any SNP within the colocalisation window was associated exclusively with inflammatory marker concentrations, tissue-specific or immune cell-specific gene expression, or cancer risk (p_1_ = 1 x 10^-4^); associated with two of these traits (p_2_ = 1 x 10^-6^); or associated with all three traits (p_3_ = 1 x 10^-7^).

### Evaluation of drug repurposing opportunities using DrugBank

For all findings showing “strong” or “suggestive evidence” in MR analysis and evidence of colocalisation, we used DrugBank to identify investigational and/or approved drugs targeting these inflammatory markers [95]. The availability of drugs targeting these markers could suggest potential for their repurposing as pharmacological agents for cancer prevention.

A step-by-step overview of GWAS selection, instrument construction, and statistical analysis stages is presented in **Figure 1**.

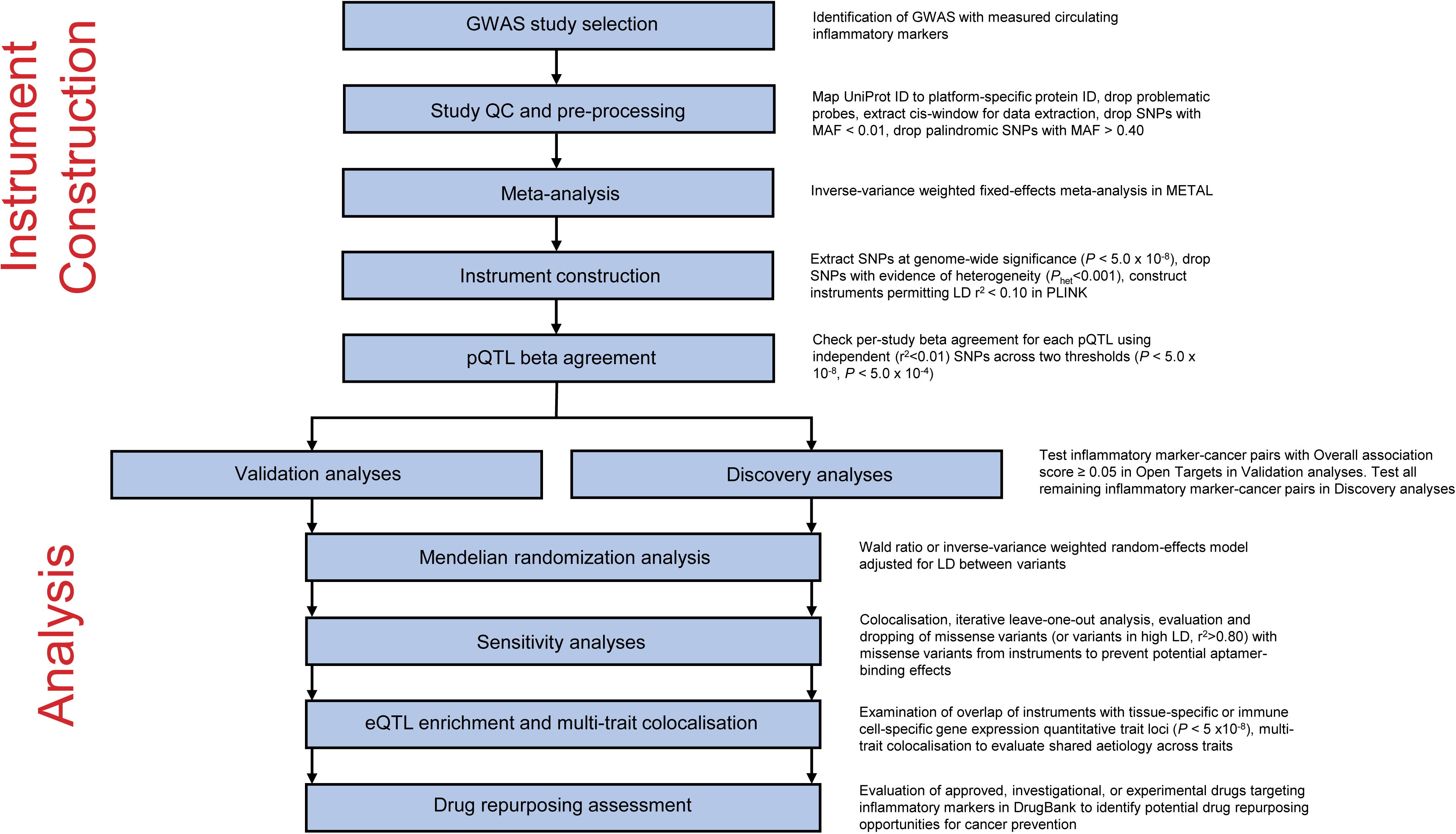

## Results

Across 66 circulating inflammatory markers, F-statistics for their instruments ranged from 30.0-7,608.0, suggesting that instruments were unlikely to suffer from weak instrument bias. Characteristics of genetic variants used to proxy circulating inflammatory markers are presented in **Supplementary Table 4**. Estimates of r^2^ and F-statistics for each marker are presented in **Supplementary Table 5**.

### Validation Mendelian randomization analyses

56 of the 66 inflammatory markers were included in validation analyses (i.e. markers with at least one cancer outcome with an overall association score ≥ 0.05 in Open Targets). In total, 260 target-cancer associations were tested. In Mendelian randomization analysis, there was suggestive evidence to support 6 inflammatory marker-cancer associations. The strongest associations were for tumour necrosis factor ligand superfamily member 10 (TRAIL) concentrations and breast cancer risk (OR 0.90 per SD increase, 95% CI 0.85-0.95, q-value=0.072), circulating macrophage migration inhibitory factor (MIF) concentrations and bladder cancer risk (OR 2.46, 95% CI 1.48-4.10, *q*-value=0.072), and interleukin-7 receptor subunit alpha concentrations and colon cancer risk (OR 0.83, 95% CI 0.74-0.93, *q*-value=0.093)(**Figure 2**). Findings for all inflammatory marker-cancer associations that employed IVW models were robust to iterative leave-one-out analysis. In colocalisation analysis, there was evidence to support shared causal variants across circulating MIF concentrations and bladder cancer risk in the *MIF* locus (PPH_4_=76.1%), but little evidence to suggest shared causal variants across 5 other inflammatory marker-cancer associations. Complete results from primary Mendelian randomization, iterative leave-one-out, and colocalisation analyses are presented in **Supplementary Tables 6-8**.

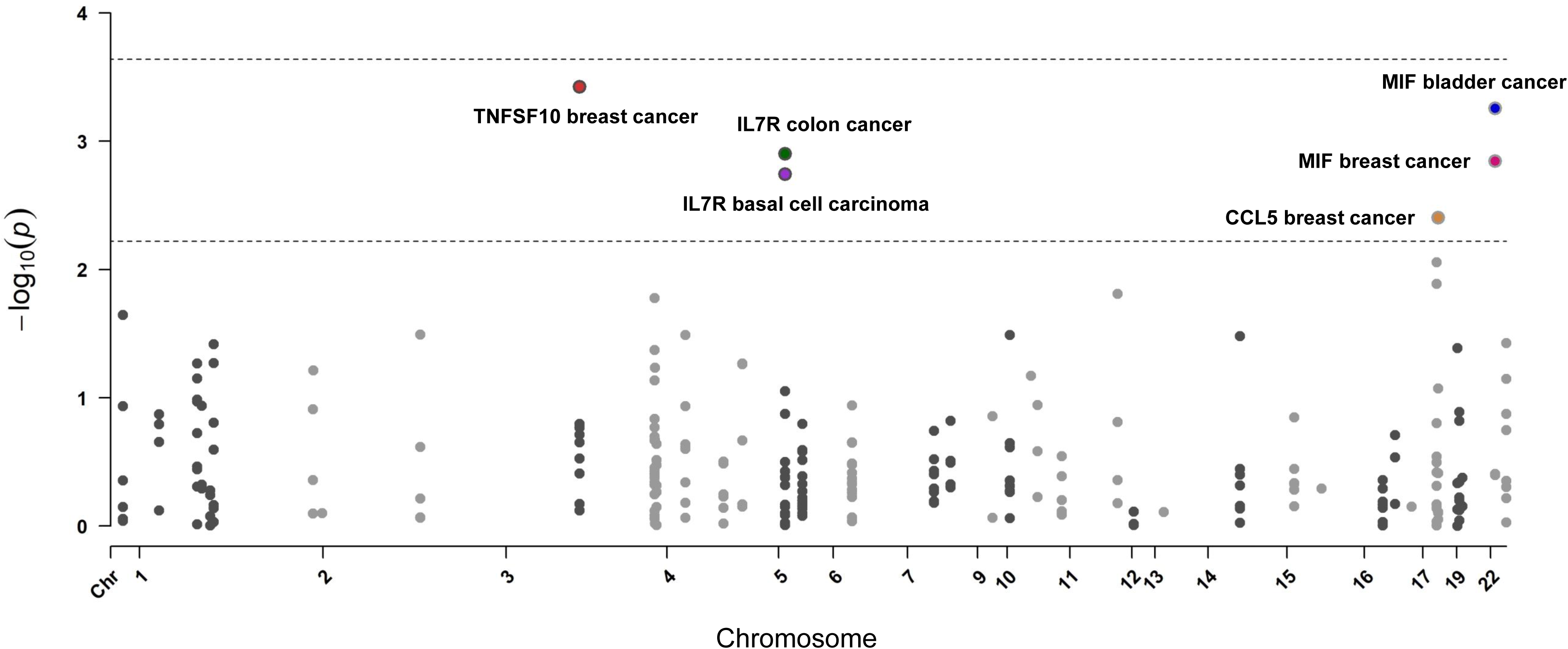

### Discovery Mendelian randomization analyses

Among the 66 circulating inflammatory markers included in Discovery analyses, there was strong or suggestive evidence for 6 inflammatory marker-cancer associations. The strongest associations were for circulating pro-adrenomedullin concentrations and breast cancer risk (OR 1.19, 95% CI 1.10-1.29; *q*-value=0.033), interleukin-23 receptor concentrations and pancreatic cancer risk (OR 1.42, 95% CI 1.20-1.69; *q*-value=0.055), prothrombin concentrations and basal cell carcinoma risk (OR 0.66, 95% CI 0.53-0.81; *q*-value=0.067), serum amyloid P component concentrations and low grade serous ovarian cancer risk (OR 2.00, 95% CI 1.41-2.83; *q*-value=0.084), and interleukin-1 receptor-like 1 concentrations and triple-negative breast cancer risk (OR 0.92, 95% CI 0.88-0.97, *q*-value =0.15)(**Figure 3**). Findings for all inflammatory marker-cancer associations that employed IVW models were robust to iterative leave-one-out analysis. In colocalisation analysis, pro-adrenomedullin and breast cancer risk (PPH_4_=94.4%), interleukin-23 receptor and pancreatic cancer risk (PPH_4_=73.9%), prothrombin and basal cell carcinoma risk (PPH_4_=81.8%), and interleukin-1 receptor-like 1 and triple-negative breast cancer risk (PPH_4_=85.6%) showed evidence of shared causal variants across traits. Complete findings from primary Mendelian randomization, iterative leave-one-out, and colocalisation analyses are presented in **Supplementary Tables 7-9**.

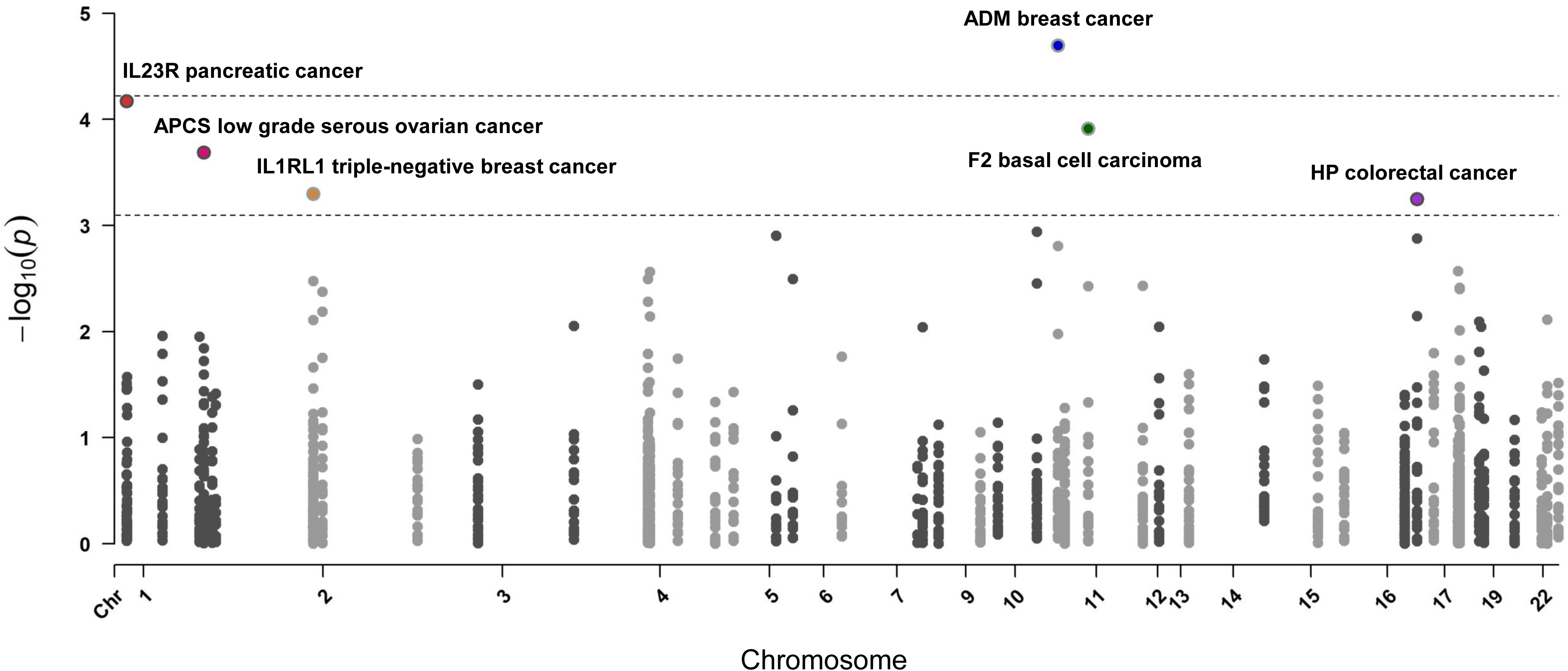

### Evaluation of aptamer or epitope binding effects of instruments

In sensitivity analyses exploring potential aptamer or epitope binding effects of instruments for Mendelian randomization findings showing evidence of colocalisation, one of two SNPs used to instrument interleukin-23 receptor concentrations (rs11581607) was in perfect LD (r^2^=1.0) with a missense variant (rs11209026). Mendelian randomization findings were consistent when dropping this SNP from the instrument and calculating a revised causal estimate (OR 1.40, 95% CI 1.10-1.78). In colocalisation analysis employing conditional analysis adjusting for rs11581607, evidence of shared causal variants across interleukin-23 receptor and pancreatic cancer risk associations persisted (PPH_4_=69.8%). The SNP used to instrument prothrombin (rs3136516) was in high LD (r^2^=0.83) with a missense variant (rs2306029). There was evidence that rs3136516 is a sQTL for *F2* (gene encoding prothrombin) in liver tissue (normalised effect size per copy of circulating prothrombin-increasing allele: 0.63, *P*=3.1 x 10^-12^). Functional annotation of SNPs used to instrument inflammatory markers along with SNPs in high LD with these variants is presented in **Supplementary Table 10**.

### Expression quantitative trait loci overlap and multi-trait colocalisation analysis

Across 5 inflammatory markers with strong or suggestive evidence of association with cancer outcomes, there was evidence that the SNP used to instrument macrophage migration inhibitory factor concentrations (rs2330634) was an eQTL for *MIF* in 35 tissue types. There was little evidence of eQTL overlap for SNPs used to instrument the 4 other inflammatory markers. In multiple trait colocalisation analysis, there were low posterior probabilities of shared causal variants across circulating macrophage migration inhibitory factor, tissue-level *MIF* expression, and bladder cancer risk (a summary of eQTL overlap and findings from multiple trait colocalisation analysis is presented in **Supplementary Table 11**).

### Genetic findings support drug repurposing opportunities in DrugBank

In DrugBank, of the 5 inflammatory markers showing evidence of a causal relationship with site-specific cancer, 3 of these markers (or related markers) are targets of approved or investigational medications. Though there are no approved medications that target the interleukin-23 receptor, three interleukin-23 inhibitors or antagonists (i.e. ustekinumab, tildrakizumab, risankizumab) have been approved to treat autoimmune conditions including moderate-to-severe plaque psoriasis [96, 97]. Several prothrombin activators or agonists have been approved for the treatment and prevention of bleeding in individuals with haemophilia A (e.g. Moroctocog alfa, Lonoctocog alfa)[98, 99]. There are several investigational and experimental medications targeting macrophage migration inhibitory factor, but no approved drugs for this target to date. There were no investigational or approved inhibitors of pro-adrenomedullin or adrenomedullin or antagonists of adrenomedullin receptor subunits (calcitonin-receptor-like receptor, receptor activity-modifying protein 2, receptor activity-modifying protein 3) and no investigational or approved medications targeting interleukin-1 receptor like-1 or the interleukin-1 receptor like-1 ligand interleukin-33 in DrugBank. A summary of investigational and approved medications targeting the 5 inflammatory markers is presented in **Supplementary Table 12**.

## Discussion

Our systematic Mendelian randomization and colocalisation analysis of 66 circulating inflammatory markers in risk of 30 cancers identified potential roles for 5 markers in risk of 5 site-specific cancers. We found suggestive evidence to implicate macrophage migration inhibitory factor concentrations in bladder cancer risk, consistent with prior reports, and identified potential novel associations between four other inflammatory markers (pro-adrenomedullin, interleukin-23 receptor, prothrombin, and interleukin-1 receptor-like 1) and risk of site-specific cancers. For 22 of 30 cancer outcomes examined (e.g. overall and subtype-specific lung, ovarian, and endometrial cancer), we found little evidence for association of circulating inflammatory markers with cancer risk despite evidence from conventional epidemiological studies suggesting roles of various markers evaluated in their aetiology [14, 18, 21, 22, 100, 101]. We also found little evidence that several putative key inflammatory mediators in cancer development (e.g. epidermal growth factor, interleukin-6 receptor, interleukin-8) were associated with cancer risk [102–104].

Some of the disagreement between our findings and those reported in the epidemiological literature could reflect the relatively limited sample size of some cancer outcomes included in our analyses (i.e. 13 of 30 outcomes were restricted to <10,000 cases). Alternatively, conflicting findings could reflect the susceptibility of conventional epidemiological studies to unmeasured or residual confounding (e.g. due to imprecisely measured confounders) and reverse causation (e.g. latent, undiagnosed cancer influencing circulating inflammatory marker concentrations). Our findings therefore do not support widespread effects of those circulating inflammatory markers evaluated on cancer risk across several anatomical sites, though we cannot rule out small to modest or potential time-varying effects of these markers or potential heterogeneity of effects across participant subgroups (e.g. long-term smokers, individuals with autoimmune conditions).

In “Validation analyses” appraising previously reported inflammatory marker-cancer risk relationships, we found suggestive evidence for an association of genetically-proxied macrophage migration inhibitory factor concentrations with bladder cancer risk. MIF is a pleiotropic inflammatory cytokine and critical upstream mediator of innate immunity [105, 106]. *In vitro* studies of human bladder tissue have supported a role of MIF in tumour cell proliferation [107]. Further, in animal models of bladder cancer, *MIF* knockout mice displayed decreased angiogenesis and invasion as compared with wild-type mice and those administered oral MIF inhibitors had decreased growth and progression of tumours as compared to controls [108, 109]. Our findings, consistent with the preclinical literature, therefore suggest a potential role of MIF as a mediator of inflammation-driven bladder cancer and support the possible utility of pharmacological MIF inhibition as preventive therapy for this cancer.

In “Discovery analyses” examining potential novel inflammatory marker-cancer pairs, we found strong evidence for an association of genetically-proxied pro-adrenomedullin concentrations with breast cancer risk. Pro-adrenomedullin undergoes proteolysis and amidation to yield adrenomedullin and proadrenomedullin N-20 terminal peptide [110]. Adrenomedullin is a potent vasodilator that is expressed in breast cancer cells, upregulated by hypoxia in these cells, and has been shown to stimulate angiogenesis and tumour proliferation [111–113]. In addition, breast cancer cells overexpressing adrenomedullin show lower levels of various apoptotic factors (e.g. Bax, Bid, caspase 8) and murine models of breast cancer with adrenomedullin overexpression have accelerated bone metastasis and lower rates of survival [113, 114].

We also found suggestive evidence for an association of genetically-proxied interleukin-23 receptor concentrations with pancreatic cancer risk. The interleukin-23 receptor pairs with the interleukin-12 receptor β-1 subunit to mediate signalling of interleukin-23, a “master regulator” of innate and adaptive immunity and promoter of inflammatory mediators in the tumour microenvironment [115, 116]. Preclinical studies have reported that interleukin-23 can promote tumour metastasis through up-regulation of angiogenic factors and that interleukin-23 receptor blockade may confer protection against tumour growth [117, 118]. In addition, serum interleukin-23 concentrations are elevated in pancreatic cancer patients as compared to controls and higher expression of interleukin-23 in tumour tissues of patients is associated with advanced clinical stage [119, 120]. Four interleukin-23 inhibitors (i.e. ustekinumab, risankizumab, guselkumab, tildrakizumab) are currently approved for the treatment of immune-mediated inflammatory diseases such as plaque psoriasis [96, 97]. To date, clinical trials of these medications have not reported links with pancreatic cancer, though sample size (≤ 1,306 participants included in trials) and duration of follow-up of studies (i.e. median 24 weeks to 2.9 years follow-up across studies) have been limited [121–125]. Though further preclinical and epidemiological work is required to validate and clarify potential mechanisms governing this effect, our findings suggesting an adverse association of genetically-proxied interleukin-23 receptor concentrations with pancreatic cancer risk provide tentative support for a potential role for interleukin-23 inhibition or antagonism as a pharmacological approach for pancreatic cancer prevention.

Finally, there was suggestive evidence to support protective associations of genetically-proxied prothrombin concentrations with basal cell carcinoma and interleukin-1 receptor-like 1 concentrations with triple-negative breast cancer risk. Prothrombin is proteolytically cleaved to form thrombin, the end-product of the coagulation cascade that converts soluble fibrinogen to a fibrin clot. Preclinical studies have reported that thrombin can induce tumour growth, metastasis, and angiogenesis and that thrombin inhibitors suppress tumour growth and metastasis in some cancer cell lines [126–128]. In our analyses, the SNP used to instrument prothrombin was in high LD with a missense variant which could influence aptamer binding and produce biased effect estimates. Though the SNP used to instrument prothrombin was also a sQTL for *F2* (gene encoding prothrombin) in liver tissue, confident causal conclusions about the direction and magnitude of an effect of genetically-proxied prothrombin concentrations on cancer risk mediated via this variant cannot be made. Interleukin-1 receptor-like 1 is the cognate receptor for interleukin-33, an epithelial-derived cytokine which has been reported to confer both pro- and anti-tumorigenic effects, depending on the tumour and cellular context, expression levels, and the nature of the inflammatory environment [129, 130]. Few studies have examined a potential role of interleukin-33 in triple-negative breast cancer risk though increased expression of this marker is associated with improved survival in triple-negative breast cancer risk [131]. The potential protective role of circulating interleukin-33 concentrations in triple-negative breast cancer risk requires further examination in future studies.

Strengths of this analysis include the comprehensive and systematic evaluation of a large number of circulating inflammatory markers, representative of different classes of inflammatory markers, in risk of 30 adult cancers. We used a dual “hypothesis-validating” and “hypothesis-generating” approach to attempt to validate previously reported inflammatory marker-cancer pairs and to identify potential novel inflammatory marker-cancer pairs. By performing a meta-analysis of 6 prior GWAS of circulating inflammatory marker concentrations we were able to generate stronger *ci*s-acting instruments for inflammatory markers examined and to develop novel instruments for some markers, increasing statistical power and the breadth of analyses performed. The use of colocalisation as a sensitivity analysis permitted us to test the robustness of Mendelian randomization findings to confounding through linkage disequilibrium. Only 5 of 12 Mendelian randomization findings with “strong” or “suggestive” evidence of association were consistent in colocalisation analyses, highlighting the importance of evaluation of shared causal variants as further support for causality of traits examined, though we cannot rule out low power influencing findings from some of these analyses.

There are several limitations to these analyses. First, the presence or absence of associations of circulating inflammatory markers may not reflect potential tissue-specific effects of these markers in cancer development. While we examined tissue-specific regulatory mechanisms underpinning effects of inflammatory markers showing strong or suggestive evidence of association with cancer risk, we did not systematically evaluate potential tissue-specific effects of genes encoding inflammatory markers across all cancer endpoints. Second, Mendelian randomization estimates represent the effect of long-term genetically-proxied inflammatory marker concentrations on cancer risk which may not correspond to the effect of pharmacological inhibition of these markers over relatively limited periods of time in clinical trials. Third, effect estimates presented assume no gene-environment or gene-gene interactions and linear and time-fixed effects of markers on cancer risk. Fourth, our analyses were restricted to cancer risk and not progression and therefore may not be informative of the utility of targeting inflammatory markers examined in the context of cancer treatment. Fifth, our analyses were performed in individuals of European ancestry and therefore the generalisability of these findings to non-European populations is unclear. Sixth, statistical power was likely limited for some rarer cancers and select cancer subtypes. Seventh, though various sensitivity analyses were employed to test robustness of our findings to potential violations of exchangeability and exclusion restriction assumptions, these assumptions are unverifiable. Eighth, we cannot rule out the possibility that the association of genetically-proxied prothrombin concentrations with basal cell carcinoma risk is driven through aptamer-binding effects given that the variant used to instrument this marker is in high LD with a missense variant. Finally, the inflammatory markers included in this analysis constitute a non-exhaustive list of inflammation-related markers which may influence cancer risk.

We found limited overlap of instruments for circulating inflammatory markers with tissue-specific or immune cell-specific eQTLs for the genes encoding these proteins which could plausibly reflect genetic effects on processes other than transcription, including protein degradation, binding, secretion, or clearance from circulation [60]. Systematic evaluation of the role of tissue-specific eQTLs for inflammation-related genes in cancer risk could provide further insight into tissue-level regulatory mechanisms influencing cancer development. There is a need to further replicate and validate findings, particularly for novel associations linking inflammatory markers to cancer risk identified in “Discovery” analyses. Future studies restricted to participant sub-groups with elevated risk of cancer (e.g. life-long smokers, individuals with chronic inflammatory conditions) could aid in identification of inflammatory markers mediating cancer risk in these groups. Finally, evaluation of the potential role of circulating inflammatory markers in cancer prognosis could inform on the possible utility of the pharmacological targeting of these markers as cancer treatment.

## Conclusion

Our systematic joint Mendelian randomization and colocalisation analysis of the effect of 66 circulating inflammatory markers in risk of 30 cancers suggests potential roles for 5 circulating inflammatory markers in risk of 5 site-specific cancers. Consistent with prior reports, we found suggestive evidence for a role of macrophage migration inhibitory factor in bladder cancer risk and strong or suggestive evidence for roles of 4 other potential novel inflammatory markers in site-specific cancer risk. Contrary to some previous conventional epidemiological studies, we found little evidence for association of circulating inflammatory markers with risk of the majority of cancer outcomes evaluated.

## Funding

JY is supported by a Cancer Research UK Population Research Postdoctoral Fellowship (C68933/A28534). SJL, KKT and RMM are supported by Cancer Research UK (C18281/A29019) programme grant (the Integrative Cancer Epidemiology Programme) (https://www.cancerresearchuk.org/). KKT is also supported by Cancer Research UK (PPRCPJT\100005) and World Cancer Research Fund (IIG_FULL_2020_022) grants. RMM is a National Institute for Health Research Senior Investigator (NIHR202411). RMM is also supported by the NIHR Bristol Biomedical Research Centre which is funded by the NIHR (BRC-1215-20011) and is a partnership between University Hospitals Bristol and Weston NHS Foundation Trust and the University of Bristol. Department of Health and Social Care disclaimer: The views expressed are those of the author(s) and not necessarily those of the NHS, the NIHR or the Department of Health and Social Care. JWR, KB, and GDS are part of the Medical Research Council Integrative Epidemiology Unit at the University of Bristol which is supported by the Medical Research Council (MC_UU_00011/1, MC_UU_00011/3, MC_UU_00011/6, and MC_UU_00011/4) and the University of Bristol (https://mrc.ukri.org/; https://www.bristol.ac.uk/).VK is funded by Academy of Finland Project 326291 and the European Union’s Horizon 2020 grant agreement no. 848158 (EarlyCause). NM is supported by the French National Cancer Institute (INCa SHSESP20, grant No. 2020-076). SSZ is supported by a National Institute for Health Research Clinical Lectureship and works in centres supported by Versus Arthritis (grant no. 21173, 21754 and 21755). INTEGRAL-ILCCO is supported by a National Institutes of Health grant (U19 CA203654) (https://www.nih.gov/). The International Lung Cancer Consortium is supported by a National Cancer Institute grant (U19CA203654) (https://www.cancer.gov/).

GECCO funding:

Genetics and Epidemiology of Colorectal Cancer Consortium (GECCO): National Cancer Institute, National Institutes of Health, U.S. Department of Health and Human Services (R01 CA059045, U01 CA164930, R01 CA244588, R01 CA201407. This research was funded in part through the NIH/NCI Cancer Center Support Grant P30 CA015704. Scientific Computing Infrastructure at Fred Hutch funded by ORIP grant S10OD028685.

ASTERISK: a Hospital Clinical Research Program (PHRC-BRD09/C) from the University Hospital Center of Nantes (CHU de Nantes) and supported by the Regional Council of Pays de la Loire, the Groupement des Entreprises Françaises dans la Lutte contre le Cancer (GEFLUC), the Association Anne de Bretagne Génétique and the Ligue Régionale Contre le Cancer (LRCC).

The ATBC Study is supported by the Intramural Research Program of the U.S. National Cancer Institute, National Institutes of Health, Department of Health and Human Services.

CLUE II funding was from the National Cancer Institute (U01 CA086308, Early Detection Research Network; P30 CA006973), National Institute on Aging (U01 AG018033), and the American Institute for Cancer Research. The content of this publication does not necessarily reflect the views or policies of the Department of Health and Human Services, nor does mention of trade names, commercial products, or organizations imply endorsement by the US government. Maryland Cancer Registry (MCR) Cancer data was provided by the Maryland Cancer Registry, Center for Cancer Prevention and Control, Maryland Department of Health, with funding from the State of Maryland and the Maryland Cigarette Restitution Fund. The collection and availability of cancer registry data is also supported by the Cooperative Agreement NU58DP006333, funded by the Centers for Disease Control and Prevention. Its contents are solely the responsibility of the authors and do not necessarily represent the official views of the Centers for Disease Control and Prevention or the Department of Health and Human Services.

ColoCare: This work was supported by the National Institutes of Health (grant numbers R01 CA189184 (Li/Ulrich), U01 CA206110 (Ulrich/Li/Siegel/Figueiredo/Colditz, 2P30CA015704-40 (Gilliland), R01 CA207371 (Ulrich/Li)), the Matthias Lackas-Foundation, the German Consortium for Translational Cancer Research, and the EU TRANSCAN initiative.

The Colon Cancer Family Registry (CCFR, www.coloncfr.org) is supported in part by funding from the National Cancer Institute (NCI), National Institutes of Health (NIH) (award U01 CA167551).

Support for case ascertainment was provided in part from the Surveillance, Epidemiology, and End Results (SEER) Program and the following U.S. state cancer registries: AZ, CO, MN, NC, NH; and by the Victoria Cancer Registry (Australia) and Ontario Cancer Registry (Canada). The CCFR Set-1 (Illumina 1M/1M-Duo) and Set-2 (Illumina Omni1-Quad) scans were supported by NIH awards U01 CA122839 and R01 CA143237 (to GC). The CCFR Set-3 (Affymetrix Axiom CORECT Set array) was supported by NIH award U19 CA148107 and R01 CA81488 (to SBG). The CCFR Set-4 (Illumina OncoArray 600K SNP array) was supported by NIH award U19 CA148107 (to SBG) and by the Center for Inherited Disease Research (CIDR), which is funded by the NIH to the Johns Hopkins University, contract number HHSN268201200008I. Additional funding for the OFCCR/ARCTIC was through award GL201-043 from the Ontario Research Fund (to BWZ), award 112746 from the Canadian Institutes of Health Research (to TJH), through a Cancer Risk Evaluation (CaRE) Program grant from the Canadian Cancer Society (to SG), and through generous support from the Ontario Ministry of Research and Innovation. The SFCCR Illumina HumanCytoSNP array was supported in part through NCI/NIH awards U01/U24 CA074794 and R01 CA076366 (to PAN). The content of this manuscript does not necessarily reflect the views or policies of the NCI, NIH or any of the collaborating centers in the Colon Cancer Family Registry (CCFR), nor does mention of trade names, commercial products, or organizations imply endorsement by the US Government, any cancer registry, or the CCFR.

COLON: The COLON study is sponsored by Wereld Kanker Onderzoek Fonds, including funds from grant 2014/1179 as part of the World Cancer Research Fund International Regular Grant Programme, by Alpe d’Huzes and the Dutch Cancer Society (UM 2012–5653, UW 2013-5927, UW2015-7946), and by TRANSCAN (JTC2012-MetaboCCC, JTC2013-FOCUS). The Nqplus study is sponsored by a ZonMW investment grant (98-10030); by PREVIEW, the project PREVention of diabetes through lifestyle intervention and population studies in Europe and around the World (PREVIEW) project which received funding from the European Union Seventh Framework Programme (FP7/2007–2013) under grant no. 312057; by funds from TI Food and Nutrition (cardiovascular health theme), a public–private partnership on precompetitive research in food and nutrition; and by FOODBALL, the Food Biomarker Alliance, a project from JPI Healthy Diet for a Healthy Life.

COLO2&3: National Institutes of Health (R01 CA060987).

Colorectal Cancer Transdisciplinary (CORECT) Study: The CORECT Study was supported by the National Cancer Institute, National Institutes of Health (NCI/NIH), U.S. Department of Health and Human Services (grant numbers U19 CA148107, R01 CA081488, P30 CA014089, R01 CA197350; P01 CA196569; R01 CA201407; R01 CA242218), National Institutes of Environmental Health Sciences, National Institutes of Health (grant number T32 ES013678) and a generous gift from Daniel and Maryann Fong.

CORSA: The CORSA study was funded by Austrian Research Funding Agency (FFG) BRIDGE (grant 829675, to Andrea Gsur), the “Herzfelder’sche Familienstiftung” (grant to Andrea Gsur) and was supported by COST Action BM1206.

CPS-II: The American Cancer Society funds the creation, maintenance, and updating of the Cancer Prevention Study-II (CPS-II) cohort. The study protocol was approved by the institutional review boards of Emory University, and those of participating registries as required.

CRCGEN: Colorectal Cancer Genetics & Genomics, Spanish study was supported by Instituto de Salud Carlos III, co-funded by FEDER funds –a way to build Europe– (grants PI14-613 and PI09-1286), Agency for Management of University and Research Grants (AGAUR) of the Catalan Government (grant 2017SGR723), Junta de Castilla y León (grant LE22A10-2), the Spanish Association Against Cancer (AECC) Scientific Foundation grant GCTRA18022MORE and the Consortium for Biomedical Research in Epidemiology and Public Health (CIBERESP), action Genrisk. Sample collection of this work was supported by the Xarxa de Bancs de Tumors de Catalunya sponsored by Pla Director d’Oncología de Catalunya (XBTC), Plataforma Biobancos PT13/0010/0013 and ICOBIOBANC, sponsored by the Catalan Institute of Oncology. We thank CERCA Programme, Generalitat de Catalunya for institutional support.

Czech Republic CCS: This work was supported by the Czech Science Foundation (21-04607X, 21-27902S), by the Grant Agency of the Ministry of Health of the Czech Republic (grants AZV NU21-07-00247 and AZV NU21-03-00145), and Charles University Research Fund (Cooperation 43-Surgical disciplines).

DACHS: This work was supported by the German Research Council (BR 1704/6-1, BR 1704/6-3, BR 1704/6-4, CH 117/1-1, HO 5117/2-1, HE 5998/2-1, KL 2354/3-1, RO 2270/8-1 and BR 1704/17-1), the Interdisciplinary Research Program of the National Center for Tumor Diseases (NCT), Germany, and the German Federal Ministry of Education and Research (01KH0404, 01ER0814, 01ER0815, 01ER1505A and 01ER1505B).

DALS: National Institutes of Health (R01 CA048998 to M. L. Slattery).

EDRN: This work is funded and supported by the NCI, EDRN Grant (U01-CA152753).

EPIC: The coordination of EPIC is financially supported by International Agency for Research on Cancer (IARC) and also by the Department of Epidemiology and Biostatistics, School of Public Health, Imperial College London which has additional infrastructure support provided by the NIHR Imperial Biomedical Research Centre (BRC). The national cohorts are supported by: Danish Cancer Society (Denmark); Ligue Contre le Cancer, Institut Gustave Roussy, Mutuelle Générale de l’Education Nationale, Institut National de la Santé et de la Recherche Médicale (INSERM) (France); German Cancer Aid, German Cancer Research Center (DKFZ), German Institute of Human Nutrition Potsdam-Rehbruecke (DIfE), Federal Ministry of Education and Research (BMBF) (Germany); Associazione Italiana per la Ricerca sul Cancro-AIRC-Italy, Compagnia di SanPaolo and National Research Council (Italy); Dutch Ministry of Public Health, Welfare and Sports (VWS), Netherlands Cancer Registry (NKR), LK Research Funds, Dutch Prevention Funds, Dutch ZON (Zorg Onderzoek Nederland), World Cancer Research Fund (WCRF), Statistics Netherlands (The Netherlands); Health Research Fund (FIS) - Instituto de Salud Carlos III (ISCIII), Regional Governments of Andalucía, Asturias, Basque Country, Murcia and Navarra, and the Catalan Institute of Oncology - ICO (Spain); Swedish Cancer Society, Swedish Research Council and and Region Skåne and Region Västerbotten (Sweden); Cancer Research UK (14136 to EPIC-Norfolk; C8221/A29017 to EPIC-Oxford), Medical Research Council (1000143 to EPIC-Norfolk; MR/M012190/1 to EPIC-Oxford). (United Kingdom).

EPICOLON: This work was supported by grants from Fondo de Investigación Sanitaria/FEDER (PI08/0024, PI08/1276, PS09/02368, P111/00219, PI11/00681, PI14/00173, PI14/00230, PI17/00509, 17/00878, PI20/00113, PI20/00226, Acción Transversal de Cáncer), Xunta de Galicia (PGIDIT07PXIB9101209PR), Ministerio de Economia y Competitividad (SAF07-64873, SAF 2010-19273, SAF2014-54453R), Fundación Científica de la Asociación Española contra el Cáncer (GCB13131592CAST), Beca Grupo de Trabajo “Oncología” AEG (Asociación Española de Gastroenterología), Fundación Privada Olga Torres, FP7 CHIBCHA Consortium, Agència de Gestió d’Ajuts Universitaris i de Recerca (AGAUR, Generalitat de Catalunya, 2014SGR135, 2014SGR255, 2017SGR21, 2017SGR653), Catalan Tumour Bank Network (Pla Director d’Oncologia, Generalitat de Catalunya), PERIS (SLT002/16/00398, Generalitat de Catalunya), CERCA Programme (Generalitat de Catalunya) and COST Action BM1206 and CA17118. CIBERehd is funded by the Instituto de Salud Carlos III.

ESTHER/VERDI. This work was supported by grants from the Baden-Württemberg Ministry of Science, Research and Arts and the German Cancer Aid. Harvard cohorts : HPFS is supported by the National Institutes of Health (P01 CA055075, UM1 CA167552, U01 CA167552, R01 CA137178, R01 CA151993, and R35 CA197735), NHS by the National Institutes of Health (P01 CA087969, UM1 CA186107, R01 CA137178, R01 CA151993, and R35 CA197735), and PHS by the National Institutes of Health (R01 CA042182).

Hawaii Adenoma Study: NCI grants R01 CA072520.

HCES-CRC: the Hwasun Cancer Epidemiology Study–Colon and Rectum Cancer (HCES-CRC; grants from Chonnam National University Hwasun Hospital, HCRI15011-1).

IWHS: This study was supported by NIH grants CA107333 (R01 grant awarded to P.J. Limburg) and HHSN261201000032C (N01 contract awarded to the University of Iowa).

Japan Public Health Center-based Prospective Study was supported by the National Cancer Center Research and Development Fund (23-A-31[toku], 26-A-2, 29-A-4, 2020-J-4) (since 2011) and a Grant-in-Aid for Cancer Research from the Ministry of Health, Labour and Welfare of Japan (from 1989 to 2010).

Kentucky: This work was supported by the following grant support: Clinical Investigator Award from Damon Runyon Cancer Research Foundation (CI-8); NCI R01CA136726.

LCCS: The Leeds Colorectal Cancer Study was funded by the Food Standards Agency and Cancer Research UK Programme Award (C588/A19167).

MCCS cohort recruitment was funded by VicHealth and Cancer Council Victoria. The MCCS was further supported by Australian NHMRC grants 509348, 209057, 251553 and 504711 and by infrastructure provided by Cancer Council Victoria. Cases and their vital status were ascertained through the Victorian Cancer Registry (VCR) and the Australian Institute of Health and Welfare (AIHW), including the National Death Index and the Australian Cancer Database. BMLynch was supported by MCRF18005 from the Victorian Cancer Agency.

MEC: National Institutes of Health (R37 CA054281, P01 CA033619, and R01 CA063464).

MECC: This work was supported by the National Institutes of Health, U.S. Department of Health and Human Services (R01 CA081488, R01 CA197350, U19 CA148107, R01 CA242218, and a generous gift from Daniel and Maryann Fong.

MSKCC: The work at Sloan Kettering in New York was supported by the Robert and Kate Niehaus Center for Inherited Cancer Genomics and the Romeo Milio Foundation. Moffitt: This work was supported by funding from the National Institutes of Health (grant numbers R01 CA189184, P30 CA076292), Florida Department of Health Bankhead-Coley Grant 09BN-13, and the University of South Florida Oehler Foundation. Moffitt contributions were supported in part by the Total Cancer Care Initiative, Collaborative Data Services Core, and Tissue Core at the H. Lee Moffitt Cancer Center & Research Institute, a National Cancer Institute-designated Comprehensive Cancer Center (grant number P30 CA076292).

NCCCS I & II: We acknowledge funding support for this project from the National Institutes of Health, R01 CA066635 and P30 DK034987.

NFCCR: This work was supported by an Interdisciplinary Health Research Team award from the Canadian Institutes of Health Research (CRT 43821); the National Institutes of Health, U.S. Department of Health and Human Serivces (U01 CA074783); and National Cancer Institute of Canada grants (18223 and 18226). The authors wish to acknowledge the contribution of Alexandre Belisle and the genotyping team of the McGill University and Génome Québec Innovation Centre, Montréal, Canada, for genotyping the Sequenom panel in the NFCCR samples. Funding was provided to Michael O. Woods by the Canadian Cancer Society Research Institute.

NSHDS: The research was supported by Biobank Sweden through funding from the Swedish Research Council (VR 2017-00650, VR 2017-01737), the Swedish Cancer Society (CAN 2017/581), Region Västerbotten (VLL-841671, VLL-833291), Knut and Alice Wallenberg Foundation (VLL-765961), and the Lion’s Cancer Research Foundation (several grants) and Insamlingsstiftelsen, both at Umeå University.

OSUMC: OCCPI funding was provided by Pelotonia and HNPCC funding was provided by the NCI (CA016058 and CA067941).

PLCO: Intramural Research Program of the Division of Cancer Epidemiology and Genetics and supported by contracts from the Division of Cancer Prevention, National Cancer Institute, NIH, DHHS. Funding was provided by National Institutes of Health (NIH), Genes, Environment and Health Initiative (GEI) Z01 CP 010200, NIH U01 HG004446, and NIH GEI U01 HG 004438.

SEARCH: The University of Cambridge has received salary support in respect of PDPP from the NHS in the East of England through the Clinical Academic Reserve. Cancer Research UK (C490/A16561); the UK National Institute for Health Research Biomedical Research Centres at the University of Cambridge.

SELECT: Research reported in this publication was supported in part by the National Cancer Institute of the National Institutes of Health under Award Numbers U10 CA037429 (CD Blanke), and UM1 CA182883 (CM Tangen/IM Thompson). The content is solely the responsibility of the authors and does not necessarily represent the official views of the National Institutes of Health.

SMS and REACH: This work was supported by the National Cancer Institute (grant P01 CA074184 to J.D.P. and P.A.N., grants R01 CA097325, R03 CA153323, and K05 CA152715 to P.A.N., and the National Center for Advancing Translational Sciences at the National Institutes of Health (grant KL2 TR000421 to A.N.B.-H.)

The Swedish Low-risk Colorectal Cancer Study: The study was supported by grants from the Swedish research council; K2015-55X-22674-01-4, K2008-55X-20157-03-3, K2006-72X-20157-01-2 and the Stockholm County Council (ALF project).

Swedish Mammography Cohort and Cohort of Swedish Men: This work is supported by the Swedish Research Council /Infrastructure grant, the Swedish Cancer Foundation, and the Karolinska Institutés Distinguished Professor Award to Alicja Wolk.

UK Biobank: This research has been conducted using the UK Biobank Resource under Application Number 8614

VITAL: National Institutes of Health (K05 CA154337).

WHI: The WHI program is funded by the National Heart, Lung, and Blood Institute, National Institutes of Health, U.S. Department of Health and Human Services through contracts HHSN268201100046C, HHSN268201100001C, HHSN268201100002C, HHSN268201100003C, HHSN268201100004C, and HHSN271201100004C.

## Disclaimer

DG is employed part-time by Novo Nordisk, unrelated to the submitted work. Where authors are identified as personnel of the International Agency for Research on Cancer / World Health Organization, the authors alone are responsible for the views expressed in this article and they do not necessarily represent the decisions, policy or views of the International Agency for Research on Cancer / World Health Organization.

## Data Availability

Summary genetic association data from GWAS meta-analyses will be made publicly available upon acceptance of this manuscript for publication.

## Acknowledgements

The authors would like to thank the participants of the individual studies contributing to the BCAC, CIMBA, CORECT, CCFR, ECAC, GECCO, INTEGRAL-ILCCO, OCAC, PANC4, and the UK Biobank study. The authors would also like to acknowledge the investigators of these consortia and studies for generating the data used for this analysis. The authors would like to acknowledge the following investigators of the OncoArray and GAME-ON1KG INTEGRAL-ILCCO analyses: Maria Teresa Landi, Victoria Stevens, Ying Wang, Demetrios Albanes, Neil Caporaso, Paul Brennan, Christopher I. Amos, Sanjay Shete, Rayjean J. Hung, Heike Bickeböller, Angela Risch, Richard Houlston, Stephen Lam, Adonina Tardon, Chu Chen, Stig E. Bojesen, Mattias Johansson, H-Erich Wichmann, David Christiani, Gadi Rennert, Susanne Arnold, John K. Field, Loic Le Marchand, Olle Melander, Hans Brunnström, Geoffrey Liu, Angeline Andrew, Lambertus A Kiemeney, Hongbing Shen, Shan Zienolddiny, Kjell Grankvist, Mikael Johansson, M. Dawn Teare, Yun-Chul Hong, Jian-Min Yuan, Philip Lazarus, Matthew B Schabath, Melinda C. Aldrich.

GECCO acknowledgments:

ASTERISK: We are very grateful to Dr. Bruno Buecher without whom this project would not have existed. We also thank all those who agreed to participate in this study, including the patients and the healthy control persons, as well as all the physicians, technicians and students.

CCFR: The Colon CFR graciously thanks the generous contributions of their study participants, dedication of study staff, and the financial support from the U.S. National Cancer Institute, without which this important registry would not exist. The authors would like to thank the study participants and staff of the Seattle Colon Cancer Family Registry and the Hormones and Colon Cancer study (CORE Studies).

CLUE II: We thank the participants of Clue II and appreciate the continued efforts of the staff at the Johns Hopkins George W. Comstock Center for Public Health Research and Prevention in the conduct of the Clue II Cohort Study. Cancer data was provided by the Maryland Cancer Registry, Center for Cancer Prevention and Control, Maryland Department of Health, with funding from the State of Maryland and the Maryland Cigarette Restitution Fund. The collection and availability of cancer registry data is also supported by the Cooperative Agreement NU58DP006333, funded by the Centers for Disease Control and Prevention. Its contents are solely the responsibility of the authors and do not necessarily represent the official views of the Centers for Disease Control and Prevention or the Department of Health and Human Services.

COLON: the authors would like to thank the COLON and NQplus investigators at Wageningen University & Research and the involved clinicians in the participating hospitals.

CORSA: We kindly thank all individuals who agreed to participate in the CORSA study. Furthermore, we thank all cooperating physicians and students and the Biobank Graz of the Medical University of Graz.

CPS-II: The authors express sincere appreciation to all Cancer Prevention Study-II participants, and to each member of the study and biospecimen management group. The authors would like to acknowledge the contribution to this study from central cancer registries supported through the Centers for Disease Control and Prevention’s National Program of Cancer Registries and cancer registries supported by the National Cancer Institute’s Surveillance Epidemiology and End Results Program. The authors assume full responsibility for all analyses and interpretation of results. The views expressed here are those of the authors and do not necessarily represent the American Cancer Society or the American Cancer Society – Cancer Action Network.

Czech Republic CCS: We are thankful to all clinicians in major hospitals in the Czech Republic, without whom the study would not be practicable. We are also sincerely grateful to all patients participating in this study.

DACHS: We thank all participants and cooperating clinicians, and everyone who provided excellent technical assistance.

EDRN: We acknowledge all contributors to the development of the resource at University of Pittsburgh School of Medicine, Department of Gastroenterology, Department of Pathology, Hepatology and Nutrition and Biomedical Informatics.

EPIC: Where authors are identified as personnel of the International Agency for Research on Cancer/World Health Organization, the authors alone are responsible for the views expressed in this article and they do not necessarily represent the decisions, policy or views of the International Agency for Research on Cancer/World Health Organization.

EPICOLON: We are sincerely grateful to all patients participating in this study who were recruited as part of the EPICOLON project. We acknowledge the Spanish National DNA Bank, Biobank of Hospital Clínic–IDIBAPS and Biobanco Vasco for the availability of the samples. The work was carried out (in part) at the Esther Koplowitz Centre, Barcelona.

Harvard cohorts: The study protocol was approved by the institutional review boards of the Brigham and Women’s Hospital and Harvard T.H. Chan School of Public Health, and those of participating registries as required. We acknowledge Channing Division of Network Medicine, Department of Medicine, Brigham and Women’s Hospital as home of the NHS. The authors would like to acknowledge the contribution to this study from central cancer registries supported through the Centers for Disease Control and Prevention’s National Program of Cancer Registries (NPCR) and/or the National Cancer Institute’s Surveillance, Epidemiology, and End Results (SEER) Program. Central registries may also be supported by state agencies, universities, and cancer centers. Participating central cancer registries include the following: Alabama, Alaska, Arizona, Arkansas, California, Colorado, Connecticut, Delaware, Florida, Georgia, Hawaii, Idaho, Indiana, Iowa, Kentucky, Louisiana, Massachusetts, Maine, Maryland, Michigan, Mississippi, Montana, Nebraska, Nevada, New Hampshire, New Jersey, New Mexico, New York, North Carolina, North Dakota, Ohio, Oklahoma, Oregon, Pennsylvania, Puerto Rico, Rhode Island, Seattle SEER Registry, South Carolina, Tennessee, Texas, Utah, Virginia, West Virginia, Wyoming. The authors assume full responsibility for analyses and interpretation of these data.

Kentucky: We would like to acknowledge the staff at the Kentucky Cancer Registry.

LCCS: We acknowledge the contributions of Jennifer Barrett, Robin Waxman, Gillian Smith and Emma Northwood in conducting this study.

NCCCS I & II: We would like to thank the study participants, and the NC Colorectal Cancer Study staff.

NSHDS investigators thank the Västerbotten Intervention Programme, the Northern Sweden MONICA study, the Biobank Research Unit at Umeå University and Biobanken Norr at Region Västerbotten for providing data and samples and acknowledge the contribution from Biobank Sweden, supported by the Swedish Research Council.

PLCO: The authors thank the PLCO Cancer Screening Trial screening center investigators and the staff from Information Management Services Inc and Westat Inc. Most importantly, we thank the study participants for their contributions that made this study possible.

Cancer incidence data have been provided by the District of Columbia Cancer Registry, Georgia Cancer Registry, Hawaii Cancer Registry, Minnesota Cancer Surveillance System, Missouri Cancer Registry, Nevada Central Cancer Registry, Pennsylvania Cancer Registry, Texas Cancer Registry, Virginia Cancer Registry, and Wisconsin Cancer Reporting System. All are supported in part by funds from the Center for Disease Control and Prevention, National Program for Central Registries, local states or by the National Cancer Institute, Surveillance, Epidemiology, and End Results program. The results reported here and the conclusions derived are the sole responsibility of the authors.

SEARCH: We thank the SEARCH team

SELECT: We thank the research and clinical staff at the sites that participated on SELECT study, without whom the trial would not have been successful. We are also grateful to the 35,533 dedicated men who participated in SELECT.

WHI: The authors thank the WHI investigators and staff for their dedication, and the study participants for making the program possible. A full listing of WHI investigators can be found at: http://www.whi.org/researchers/Documents%20%20Write%20a%20Paper/WHI%20Investigator%20Short%20List.pdf

## Data availability statement

We obtained summary genetic association data on breast cancer risk from the Breast Cancer Association Consortium (https://bcac.ccge.medschl.cam.ac.uk/), ovarian cancer risk from the Ovarian Cancer Association Consortium (https://ocac.ccge.medschl.cam.ac.uk/), endometrial cancer risk from the Endometrial Cancer Association Consortium (https://www.ebi.ac.uk/gwas/publications/30093612#study_panel), non-Hodgkin lymphoma risk from Burrows et al. (10.5523/bris.aed0u12w0ede20olb0m77p4b9), and basal cell carcinoma risk from Adolphe et al (https://www.ebi.ac.uk/gwas/publications/33549134#study_panel). Approval was received to use restricted summary genetic association data from GECCO, INTEGRAL ILCCO, and PRACTICAL consortia after submitting a proposal to access this data. Summary genetic association data from these consortia can be accessed by contacting GECCO (, INTEGRAL ILCCO (https://ilcco.iarc.fr/), and PRACTICAL. Approval was also received to use restricted summary genetic association data on pancreatic cancer risk via dbGaP release phs000206.v5.p3. To enquire about gaining access to summary genetic association data for renal and head and neck cancer risk, contact. To enquire about gaining access to summary genetic association data for bladder cancer risk, contact.

